# Operational insights for larval source management programs: An exploratory study of *Anopheles* breeding habitat dynamics across urban wards in Ibadan, Nigeria

**DOI:** 10.64898/2026.07.16.26358299

**Authors:** Eniola A. Bamgboye, Monsuru A. Adeleke, Olabanji A. Surakat, Laurette Mhlanga, Kamilu. A. Fasasi, Akinlabi M. Rufai, K.O.K Popoola, Umar M. Aminu, Nnenna Ogbulafor, Ifeoma D. Ozodiegwu

## Abstract

Larval source management (LSM) is a complementary malaria control intervention, yet evidence to guide context-specific implementation remains limited. Nigeria’s recent national commitment to LSM scale-up makes the need for operational evidence particularly urgent. Informal settlements embedded within wards of differing dominant settlement archetypes may present distinct *Anopheles* larval habitat profiles with implications for how LSM strategies should be tailored. We evaluated *Anopheles* larval habitats within informal settlement areas across wards with contrasting settlement archetypes in Ibadan metropolis, Nigeria, to inform targeted larval source management.

Potential breeding habitats were surveyed in dry and wet seasons within informal settlement areas across three wards — Olopomewa, Challenge, and Agugu — representing formal, informal, and slum settlement-dominant archetypes respectively. Habitats were characterized and assessed fo*r Anopheles* larval presence. Pareto analysis identified habitats accounting for 80% of larval abundance. Breeding habitat density per km² was estimated using a simulated pathway technique. Associations between mosquito dispersal scale and household malaria infections identified through Rapid Diagnostic Testing were evaluated using kernel-based distance-decay weighting. Environmental drivers of habitat suitability were modeled in MaxEnt.

Of 420 potential breeding habitats identified, 31 (7.4%) contained Anopheles larvae, predominantly during the wet season (26, 83.9%). Puddles, dug wells, drainages/gutters/ditches and canals accounted for 80% of site-level larval abundance when standardized by sampling effort. Larval and breeding habitat density were highest in Agugu, the slum-dominant ward, across both seasons. Modeled mosquito dispersal scale showed best fit at 30–32m in Challenge (OR 1.41, 95% CI: 1.05–1.89) during the wet season and 16–18m in Agugu (OR 1.29, 95% CI: 1.04–1.60) during the dry season. Habitat suitability in Agugu was higher farther from large water bodies and in areas with higher population density and positive Normalized Difference Water Index values. In Challenge, suitability was higher in areas with lower nighttime light levels, positive Normalized Difference Water Index values, and negative Normalized Difference Moisture Index values.

Further studies incorporating multiple wards across diverse urban settings are needed to determine whether differences in larval ecology between settlement archetypes provide a reliable basis for planning larval source management.

## Introduction

Vector control interventions have played a prominent role in reducing malaria burden in sub-Saharan African countries, particularly in Nigeria, through the deployment of insecticide-treated nets (ITNs) and indoor residual spraying in communities at risk. [1] Yet these conventional measures face growing limitations in urban settings, where limited ITN use, emerging evidence of rising outdoor malaria transmission, and heterogeneous transmission patterns complicate control efforts. [2–3] These challenges have increasingly led to advocacy for supplementary interventions such as larval source management (LSM), which offers a complementary approach to reducing malaria transmission in urban environments.

LSM prevents *Anopheles* mosquitoes from reaching adulthood by targeting their aquatic habitats through environmental management, larviciding, and biological control. Environmental management includes interventions such as drainage improvements, filling or leveling road depressions, and managing standing water, measures that address the ecological conditions that create and sustain mosquito breeding sites. Larviciding involves the repeated application of biochemical agents to water bodies to kill *Anopheles* larvae during their developmental stages. Biological control uses natural predators such as larvivorous fish, copepods, or predatory insects to suppress larval populations in suitable aquatic habitats, providing an environmentally friendly and potentially self-sustaining complement to other LSM strategies. [4]

The World Health Organization (WHO) recommends larviciding in sub-Saharan Africa for urban areas where breeding sites are few, fixed, and readily identifiable — that is, where sites are limited in number, stable in location, and easy to locate for repeated treatment. However, larviciding is resource-intensive, requiring frequent reapplication due to short residual efficacy and extensive logistical planning where sites are widespread. Similarly, environmental management is cost-intensive, requiring sustained staffing, frequent monitoring, and in some cases infrastructural upgrades. [5] The need for context-specific evidence to guide the strategic deployment of LSM in a manner that minimizes costs is therefore considerable — and in urban Nigeria, particularly important.

This gap has become increasingly urgent in the context of Nigeria’s evolving malaria control landscape. The National Malaria Elimination Program (NMEP) recently revised its ITN distribution strategy due to funding constraints, reprioritizing future campaigns toward high malaria risk communities. [6] As high-risk designations under the revised strategy fall predominantly in rural communities, many urban areas risk exclusion from forthcoming ITN campaigns, making LSM an increasingly important strategy for urban malaria control. In recognition of this need, the Nigerian National Council of Health has adopted LSM, particularly larviciding, as a key vector control strategy and approved a larviciding feasibility pilot across six states: Lagos, Ondo, Ekiti, Rivers, Borno, and Abia. Generating the operational evidence needed to support this pilot, and subsequent LSM rollouts, is therefore a pressing priority.

Malaria transmission in urban areas is heterogeneous, and urban environments are typically characterized by rapid population growth and the expansion of informal and slum settlements, which often exhibit poorly developed basic services and infrastructure. These conditions create numerous artificial and transient aquatic habitats. [7] Critically, the density and character of larval habitat formation may be shaped by the surrounding built environment — including the extent of paved surfaces, drainage networks, solid waste management, and population density — meaning that informal settlements embedded within formally developed wards may present different habitat profiles than equivalent settlements within predominantly slum or informal wards. This distinction has received little attention in the literature but may have direct relevance for how LSM strategies are planned and targeted.

Existing studies from urban Nigeria have documented the diversity of *Anopheles* larval habitats, including ground pools, rice fields, discarded tires, containers, and gutters or canals [8–10], and have reported higher breeding site numbers and larval densities during the wet season in urban slums. [11–12] However, few studies have translated these observations into ward-level operational evidence, particularly given the settlement heterogeneity that characterizes most urban wards. Key questions remain unanswered: whether the same LSM strategies should be applied in informal settlements located in wards with different dominant settlement archetypes, and whether habitat productivity, spatial configuration, and environmental drivers vary systematically across these contexts.

Through this exploratory study, we integrate entomological and malaria infection data collected across selected urban wards in Ibadan to offer preliminary, hypothesis-generating evidence on *Anopheles* larval habitat profiles and their connections to malaria risk within informal settlements across wards of differing settlement archetypes. Specifically, we aimed to quantify entomological differences between informal settlements in wards classified as formal-, informal-, and slum-dominated by: (1) characterizing potential *Anopheles* larval habitats and estimating larval densities; (2) assessing associations between habitat positivity and seasonal characteristics; (3) estimating site-level larval productivity; (4) estimating breeding habitat density and quantifying distance-dependent spatial relationships between household malaria risk and *Anopheles* larval habitats. In our final analysis, we predict larval habitat suitability at the ward level using data from these informal settlements to identify the strongest environmental predictors for these ward archetypes.

## Methods

### Study site description

The Ibadan metropolitan area is in Oyo state, southwestern Nigeria, and is divided administratively into five Local Government Areas (LGAs), which are further subdivided into 59 wards. Ibadan is one of the largest cities in Africa by geographical area and has a mean population density of 12,665 individuals per square kilometer (SD: 5,802). [13] The city lies at an altitude of approximately 150–275 m above sea level within the savanna ecological zone (7°23′N, 3°54′E). [14]. Its landscape is shaped by rapid and ongoing urbanization comprising of vegetation that is a mosaic of secondary forest, savanna, and cultivated land interspersed with wetlands and urban agricultural plots. [14] Ibadan has a tropical climate characterized by two rainy seasons (April–July and September–October), with mean annual rainfall of approximately 1,200–1,400 mm. [14]

### Study design and context

This entomological study was conducted as part of a larger field study assessing the burden and determinants of malaria transmission in Ibadan, Nigeria. The overall aim of the parent study was to estimate malaria prevalence at the ward level, the smallest administrative unit in Nigeria, identify drivers of transmission and characterize entomological indices across wards to inform the tailoring of interventions in urban settings. [15] The entomological component described here was cross-sectional, and malaria infection status among participants was determined using Rapid Diagnostic Tests administered during the epidemiological (household survey) component of the study.

The Ibadan metropolitan area was selected for this study in collaboration with the NMEP. Its urban character, large population size, and the substantial historical investment in malaria control activities in the region make it a strategic location for identifying potential programmatic efficiencies. The selection process for the wards included in the entomology and epidemiological study were based on a model-based clustering algorithm [16] and has been previously described. [15]

### Ward-Settlement Classification

The parent study included four urban wards consisting of Enumeration Areas (EAs). EAs are clusters of households aggregated for the purposes of census data collection. For the entomological component, three wards — Olopomewa, Challenge, and Agugu — were purposively selected. Olopomewa is situated in Ibadan Northwest Local Government (LGA), Challenge in Oluyole LGA and Agugu is located in Ona Ara LGA. In each ward, EAs were used as the spatial units for identifying and classifying settlements within each selected ward. Each EA was classified as formal, informal, or slum using a settlement classification checklist developed through a multi-stakeholder dialogue process described by Ozodiegwu et al. [15]. In summary, formal-classified settlements were areas with painted building structures constructed with high-quality materials, fences, a working drainage system, water storage tank, tarred road networks and structured housing layouts with street names and house numbers. Neither informal-nor slum-classified settlements have any of these features. Informal settlements were differentiated from slums by the presence of economic activity such as shops and poorly functioning drainage systems while slums, an extreme form of informal settlements, had no drainage system in place and were characterized by open dumpsites, a generally unkept environment and the presence of abandoned houses. Olopomewa’s settlement classification revealed a combination of formal and informal settlements, with a higher percentage (56.7%) of EAs classified as formal settlements. Settlement classification in Challenge indicated a preponderance of informal settlements (91%) while the rest were formal settlements. Agugu was characterized predominantly by slum settlements (58.8%), with informal settlements accounting for the rest of the EAs. The distribution of the settlement composition by ward is presented in S1 Appendix.

### Entomological and Epidemiological studies

The entomological and epidemiological studies are described below in the order in which they were conducted.

#### a. Dry season larval habitat sampling

Larval habitat sampling in the dry season was conducted in each of the three wards selected from the parent study between January and March 2023. Enumeration areas with predominantly informal settlements were purposively selected as study sites. Five informal-classified EAs were selected each in Olopomewa and Challenge, while in Agugu, six informal-classified and one slum-classified EA were selected

#### b. Epidemiological study

The epidemiological study was conducted in the same wards as the entomological surveys during the wet (August–December 2023) and dry (January–May 2024) seasons. Using the list of classified EAs as the sampling frame, an average of 30 EAs were selected within each ward by simple random sampling using randomly generated numbers in Microsoft Excel, with representation of formal, informal, and slum settlements proportionate to their ward-level distribution. Within each selected EA, households were enumerated to create a sampling frame, from which 35-50 households per EA were randomly selected, taking into account building structure and the number of households per structure. In buildings with multiple households, particularly in the informal and slum settlement predominated wards – Challenge and Agugu, a maximum of three households per structure were selected. Full details of the sample size estimation and sampling procedures have been described in the study protocol.[15]

A total of 3,129 and 3,709 households were sampled in the wet and dry seasons, respectively, and 7,802 and 7,790 household members, respectively, were tested for malaria using rapid diagnostic tests (RDTs). Within each household, a maximum of five consenting individuals across predefined age groups were tested. Testing details are also extensively described within the study protocol [15]. Only malaria testing data from households located within the convex hull (smallest convex polygon that completely encloses a set of points, in this case the potential breeding habitats) in each of the wards during the entomological study were included in the final analysis.

#### c. Wet season entomological study

From the three wet season study wards, only two were selected due to budget constraints: Agugu (slum-predominated ward) and Challenge (informal-settlement-predominated ward). Anopheles larval habitat prospection was conducted between mid-July and early August 2024, thrice weekly. In Agugu, larval prospection covered nine EAs, comprising five slum-classified and four informal EAs, while in Challenge, prospection spanned 27 EAs, including 25 informal and two formal EAs. Fig 1 provides a schematic diagram of the site-selection process for both epidemiological and entomological surveys.

**Fig 1.**
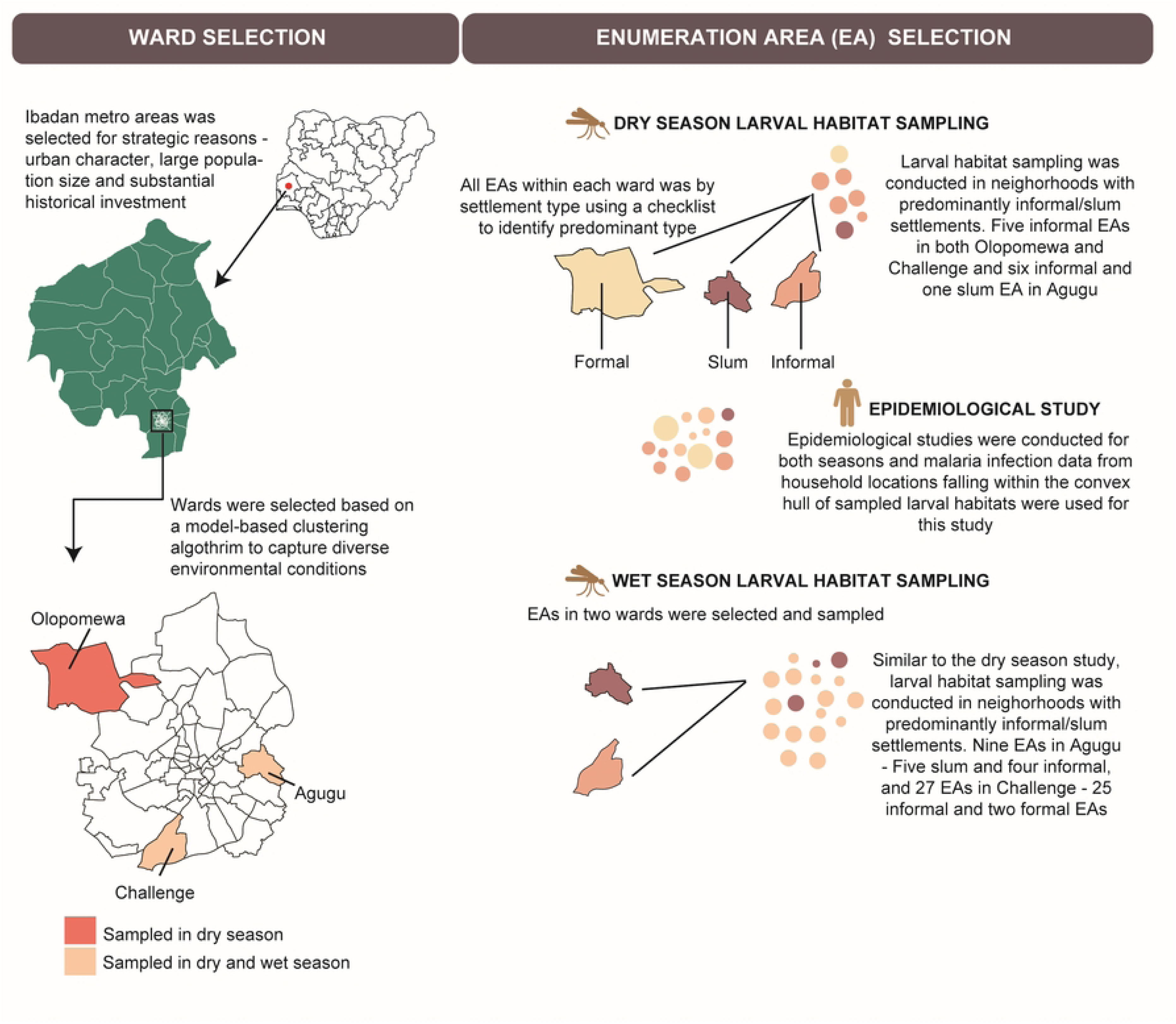
Schematic diagram of the site selection process for the larval habitat sampling and household survey data used for this study

### Data Collection

#### Breeding habitat identification

In this study, a potential *Anopheles* larval habitat was defined as any accessible natural or artificial water-holding site with standing or slow-moving water that could support the development of immature *Anopheles* mosquitoes, whether or not larvae were present at the time of inspection. [4] Potential *Anopheles* larval breeding habitats were identified using a household-centered approach, beginning at PSC-selected households and extending into surrounding accessible areas. In each selected neighborhood local community guides were recruited based on their familiarity with the neighborhood layouts and road networks. Guides were briefed on the operational criteria for identifying potential breeding habitats and accompanied entomology field teams to support navigation along access routes, drainage lines, footpaths, and open spaces. Final habitat identification, classification, and larval sampling were conducted by trained field personnel. Each breeding habitat identified during the survey was assigned a unique identification code and classified visually based on its operational definition (see S1 Table for operational definition)

#### Breeding habitat characterization

Identified potential breeding habitats were grouped based on their stability and duration to retain water into two standard categories: permanent and temporary. [17] *Permanent breeding habitats* were defined as habitats capable of holding water for extended periods, often persisting throughout the year. They include perennial drainages, ditches, canals, streams, and gutters. *Temporary breeding habitats* were described as those that result from typically retaining water for shorter durations. These include artificial containers, discarded tyres, tyre tracks, abandoned wells, potholes, and puddles. Figure 2 presents an amalgamation of photographs of these different habitat types.

**Fig 2.**
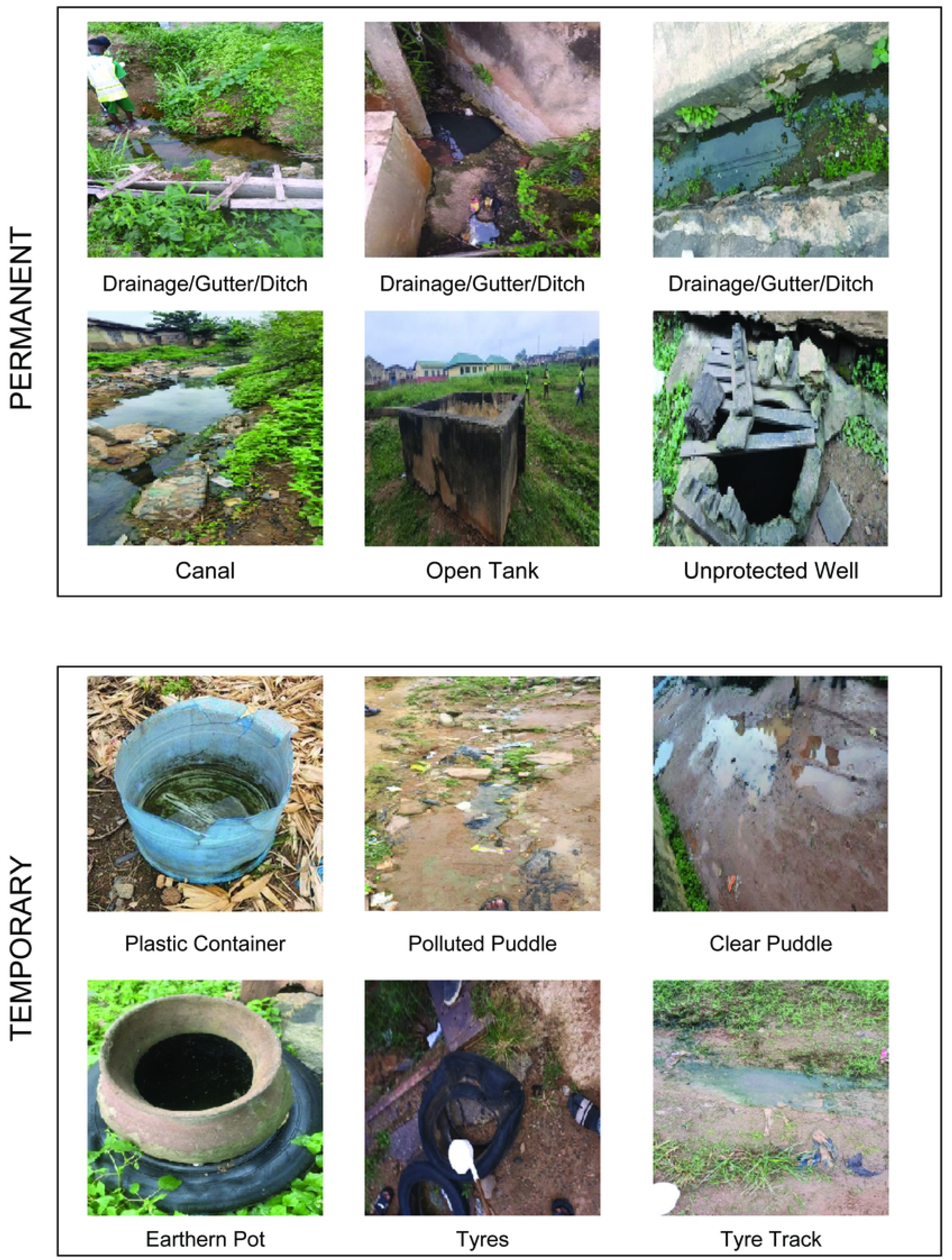
Pictures of some of the prospected habitats across the study locations

#### Breeding habitat physical and physico-chemical characteristics

For each potential breeding *habitat* identified, we documented key physical features—such as water source, water quality (clear or polluted), presence of vegetation, and exposure to sunlight using an observational checklist while the pH and temperature were measured in situ using calibrated handheld instruments. The findings were systematically collected using an Android-based Kobo Collect application. Furthermore, the geographic coordinates and photographic records of each site were captured and stored on Android devices in real time.

#### Mosquito larval sampling

Larval sampling was conducted in the early hours of the day, between 06:00 and 09:00 hours, following standard entomological procedures. At each identified potential breeding habitat, larval collections were performed using a standard 350 mL dipper in accordance with World Health Organization (WHO) guidelines [18]. The number of dips per habitat was based on habitat ￼size and accessibility￼, with 10–20 dips collected per breeding habitat. At each potential *habitat*, the total number of dips and larvae collected were recorded on the Android devices using the Kobo Collect application. Sampling locations varied throughout the study period, with surveys conducted once per week for three months during the dry season and three times per week for four consecutive weeks during the wet season. During each survey, newly observed potential larval habitats were identified and sampled.

#### Molecular identification of collected larvae

Collected larvae were morphologically identified in the field to distinguish *Anopheles* from other mosquito species based on standard larval identification keys. [19–20]. The larvae were reared to adults in the laboratory and then subjected to molecular analysis for species identification using the procedure described by Wilkin et al [21]

### Data Analysis

#### Characteristics of breeding habitat

Descriptive analyses were conducted to characterize prospected breeding *habitat* and those positive for *Anopheles* larvae, overall and by season and wards. Single variable logistic models were also used to examine associations between habitat positivity and breeding habitat type (permanent/temporary) and season. All statistical analyses were conducted at a 5% significance level (p < 0.05).

We also computed larval density per habitat using the formula below.

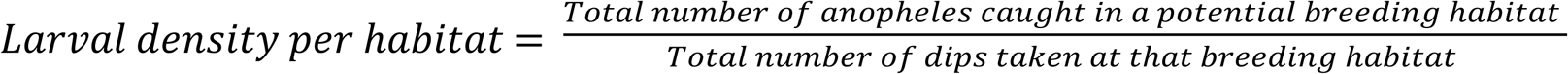

Mean larval densities were estimated among larval-positive breeding *habitat* categories by ward and season. Differences in average larval density by season and ward were explored using the Kruskal-Walli’s rank sum test due to the small sample size.

To identify priority Anopheles-positive habitats in the wet season, pareto analysis was conducted to determine breeding habitat types contributing the most to overall larval abundance. Larval densities from all prospected breeding habitats of the same type (e.g., puddles, drainages/gutters/ditches, tyres) were averaged and used as a representative measure of larval density for each habitat type, to account for the number of dips during collection.

These average larval densities were then ranked in descending order of contribution and expressed as cumulative percentages. This approach allowed identification of high-yielding breeding habitat types that contributed most to overall larval abundance, following the 80/20 principle. Statistical analyses and visualizations were carried out using R software.

#### Estimation of Anopheles larva-positive breeding habitat density

To estimate the density of Anopheles larval habitats within each survey location, while accounting for uncertainty in the areas covered during field surveys, we applied a simulated pathway sampling approach based on distance-sampling principles. This method assumes that the simulated pathways approximate an unbiased sample of the study area, that breeding habitat detected along pathways are independent events, that all breeding habitats within the effective search buffer are equally detectable and that extrapolation from sampled areas to entire study area is linear. [22]

Analysis was conducted separately for dry and wet season entomological data. We began by utilizing our georeferenced dataset of potential breeding habitat locations, which included both positive and negative *Anopheles* larval habitat across the ward-level study locations, from which two fixed points — representing a start and end breeding habitat — were identified. Next, a pathway sampling strategy was implemented by generating 1000 simulated pathways between these fixed points. At each iteration, 80% of the remaining potential breeding habitat were randomly sampled without replacement and combined with the fixed start and end breeding habitat to form a unique pathway.

Each simulated pathway was then converted to spatial lines for distance estimation, and its total length was computed as the sum of geodesic distances from consecutive points using Vincenty’s formula for geodesic distance [23]

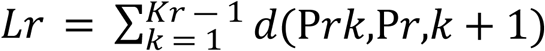

where L_r_ is the total length of simulated pathway r, K_r_ is the number of points in that pathway, P_rk_ is the (k)-^th^ point along pathway, r, and (d (P_rk_, P_r_, k+1) is the Vincenty geodesic distance between consecutive points. The sampled area for each pathway was then estimated by applying the effective strip width around the pathway:

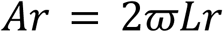

where A_r__ is the sampled area for pathway r, w is the maximum detection distance from the pathway, and L_r_ is the pathway length. The factor of 2 reflects sampling on both sides of the pathway.

Subsequently, the number of potential breeding *habitat* positive for *Anopheles* larvae within each pathway was tallied. Finally, breeding habitat density per square kilometer was estimated as the ratio of positive habitat to the sampled area. [22] Ward-level study location density estimates were summarised across the 1000 simulated pathways using the mean and uncertainty intervals from the simulated distribution:

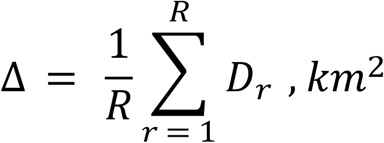

where D_r_, km^2^ is the estimated density of Anopheles-positive breeding habitat per km² for simulated pathway, r and R = 1000

An example of the output from the simulation of pathways to estimate breeding habitat density in Agugu survey location during the wet season is shown in S2 Appendix.

#### Multiscale associations between breeding habitat and household malaria positivity

To quantify household level exposure to anopheles larvae breeding habitat, we implemented a kernel-based weighting procedure informed by established distance decay formulations in spatial epidemiology capturing spatial relationships between observations according to their geographical proximity [24].

From the dry and wet season household surveys conducted as part of the main study, we extracted the geocoordinates of households within the EAs visited in each ward and overlaid these with georeferenced *Anopheles* larval habitats within the same study area. We assumed that each prospected larval habitat had the potential to produce adult mosquitoes capable of transmitting malaria. In each study location, convex hulls were generated around identified larval breeding habitats to delineate the spatial extent of habitat occurrence. Households falling within these convex hulls were extracted from the corresponding EAs and included in the analysis, regardless of settlement classification, to represent households located in areas of potential exposure to nearby breeding habitats. Household and larval habitat coordinates were then projected to UTM Zone 31N to enable accurate distance and area calculations.

Furthermore, household-level malaria status was obtained from the household survey and dichotomized as positive (if at least one household member tested positive for malaria using a rapid diagnostic test) or negative (if no household member tested positive). For each household, the Euclidean distance to all nearby larval habitat was then calculated, generating a household–larval distance matrix and these were converted into exponentially decaying weights as a function of an assumed mosquito dispersal scale (λ) using:

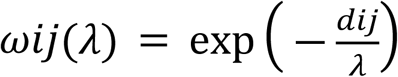

where *ω*ij is the distance-decay weight between household (*i*) and larval habitat (*j*), dij is the Euclidean distance in metres between household *(i)* and larval habitat *(j)* and ￼￼

For each candidate dispersal scale, *λ*, household exposure was defined as the mean of the distance-decayed weights across all larval habitats:

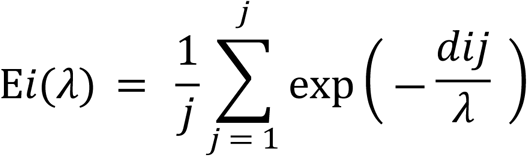

where, E*_i_*(*λ*) is the exposure score for household (*i*) at dispersal scale λ, *J* is the total number of mapped larval habitats, dij is the Euclidean distance in metres between household *(i)* and larval habitat *(j)*

Exposure values were standardised to a mean of zero and standard deviation of one before modelling. e=Effect estimates represent the odds ratio for household malaria positivity per 1 SD increase in distance-decayed larval habitat exposure. We then evaluated dispersal scale, *λ*, values from 2 m to 500 m, in 2 m increments. For each range of values, we fitted a logistic regression model with household malaria positivity as the outcome and standardised larval habitat exposure as the predictor. Model fit was compared using Akaike’s information criterion, and the *λ* with the lowest AIC was selected as the best-fitting spatial scale of association. [25] We report the selected *λ*, odds ratio and 95% confidence interval for each season and ward as applicable.

Results are presented for Agugu (slum-predominant ward) and Challenge (informal-settlement predominant ward) in the dry and wet seasons while estimates for Olopomewa, the formal-settlement-predominant ward, are not presented because sparse data produced an unstable coefficient with a non-finite confidence interval.

#### Identification of environmental predictors

Habitat suitability modeling was conducted using MaxEnt (version 3.4.3), a machine-learning approach that estimates relative habitat suitability (probability of occurrence) by comparing environmental predictors at confirmed larval-positive locations (presence points) with predictors at a sample of locations drawn from across the study area to represent available environmental conditions.

In MaxEnt, the relative suitability of a location, (*x*), is estimated as

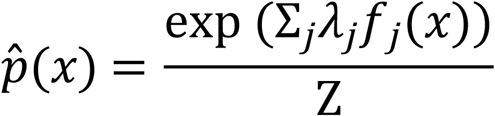

*where x represents a location, f_j_*(*x*) *are environmental predictor features; λ_j_ are fitted weights or coefficients and *Z* is a normalising constant ensuring the distribution sums to 1*

This approach is well suited to studies with relatively few location points (confirmed larval-positive sites), as was the case in our study. [26]

The geo-coordinates of the larval habitats where we found larvae in both seasons were compiled as location (presence) points. However, the dry-season dataset comprised only two and three confirmed presence points in Agugu and Olopomewa survey locations respectively, precluding robust species distribution modelling and preventing the development of season-specific models.

To address this, we developed three models: one combining dry- and wet-season observations for Agugu survey locations, and separate wet-season models for Agugu survey locations and Challenge survey locations. A combined model was not developed for Challenge survey locations because there were no positive habitats during the dry season larval prospection.

For our suitability model, we included environmental variables, which have been documented to create optimal condition for *Anopheles* larvae survival[9, 27] We incorporated Enhanced Vegetation Index (EVI) [28], Normalized Difference Moisture Index (NDMI) [28], Normalized Difference Water Index (NDWI) [28], land surface temperature [29], population density [30], building morphology [31], distance to water bodies [32], land use pattern [33] and nighttime lights [34], derived from their respective source datasets and acquisition years. (S2 Table)

Climatic variables (precipitation, atmospheric temperature, diurnal temperature range, and rainfall) were initially considered but these were excluded from the analysis due to lack of variability at the ward level resolution of our study area.

To account for lagged vegetation effects, monthly values of EVI, NDMI, and NDVI were extracted for the two months preceding the survey (January 2023–February 2023 for the dry season; May–June 2024 for the wet season), as well as last month of data collection capturing the peak of the dry and wet season (March 2023 and July 2024, respectively). We then conducted a sensitivity analysis by fitting three separate models using average vegetation indices over (i) the three-month period, (ii) the two months preceding the survey, and (iii) the last month of data collection alone.

Multicollinearity among predictors was assessed using pairwise correlation analysis; variables with correlation coefficients greater than 0·7 were excluded from analysis. The results of the correlation are presented in S3 Appendix for Agugu and S4 Appendix for Challenge. All predictor layers were clipped to the study area, resampled to a uniform spatial resolution of 30 m and projected to the same coordinate system (EPSG:32631) to ensure comparability. A spatial resolution of 30m was chosen due to the granular extent of our study area, that is ward-level, to ensure coarseness and better predictability.

Model accuracy was evaluated using the area under the receiver operating characteristic curve (AUC). Variable importance was assessed using permutation importance and jackknife tests as implemented in MaxEnt.

### Statement of Ethics

Ethical approval for this study was granted by Nigeria’s National Health Research Ethics Committee (Approval Number: NHREC/01/01/2007–10/10/05/2022), Health Research Ethics Committees of Oyo State Ministry of Health (Reference number: AD 13/479/44421A), University of Ibadan Ethics Committee (Registration number: NHREC/05/01/2008a), UNIOSUN Ethical and Research Committee(ARIP/UHRC/01); Northwestern University (IRB ID: STU00217380-MOD0001) and retrospectively from Loyola University Chicago (LU Number: 218531) when the lead and senior authors changed institutions.

## Results

### Characterization of sampled Anopheles larval habitats

#### Potential breeding habitats prospected and sampled location description

A total of 420 potential breeding habitats were identified and surveyed in the selected wards, with 151 (36.0%) habitats assessed during the dry season and 269 (64.0%) during the wet season. The number of households associated with each sampled EA where larval habitat prospecting was carried out varied by ward and season. In the dry season, prospection in Olopomewa occurred in five informal EAs, where a total of 61 households resided, and the number of households per EA ranged from 1–30 (median: 12, IQR: 2–16). In Challenge, where prospecting was conducted around five informal EAs, there were a total of 35 households, and the number of households per EA ranged from 1–19 (median: 1, IQR: 1–13). In Agugu, with six informal EAs and one slum EA, there was a total of 127 households, and the number of households per EA ranged from 2–38 (median: 10, IQR: 7–31.5). During the wet season, in Challenge, which covered 25 informal and two formal EAs, there was a total of 344 households in the surveyed location, and the number of households per EA ranged from 2–31 (median: 10, IQR: 6–19). In Agugu, five slum and four informal EAs were covered, and the number of households overall was 218, while per EA it ranged from 12–40 (median: 27, IQR: 18–28).

The EAs associated with the prospected breeding sites and households totaled an area of 0.45 km² during the dry season study in Olopomewa. The total area associated with the EAs was 0.25 km² in the dry season and 0.52 km² in the wet season for Agugu, and 0.09 km² in the dry season and 1.90 km² in the wet season for Challenge.

Both permanent and temporary breeding sites were equally found. Permanent breeding habitats consisted of drainages/gutters/ditches (192, 45.7%), and canals (18, 4.3%) while temporary breeding habitats included tyres (98, 23.3%), puddles (34, 8.1%), artificial containers (33, 7.9%), dug wells (23, 5.5%), open tanks (11, 2.6%), tyre tracks (5, 1.2%) and refuse/sewage (6, 1.4%) (Fig 3A). The proportion of permanent breeding habitats was relatively consistent across seasons, accounting for 51.4% of habitats in the dry season and 48.6% in the wet season (Fig 3B). However, a significantly higher proportion of temporary breeding habitats were observed during the wet season (79.5%) compared to the dry season (20.5%) (p < 0.05). The seasonal breakdown of the types of permanent and temporary breeding habitat types is presented in S1 Fig, panels A and B.

**Fig 3.**
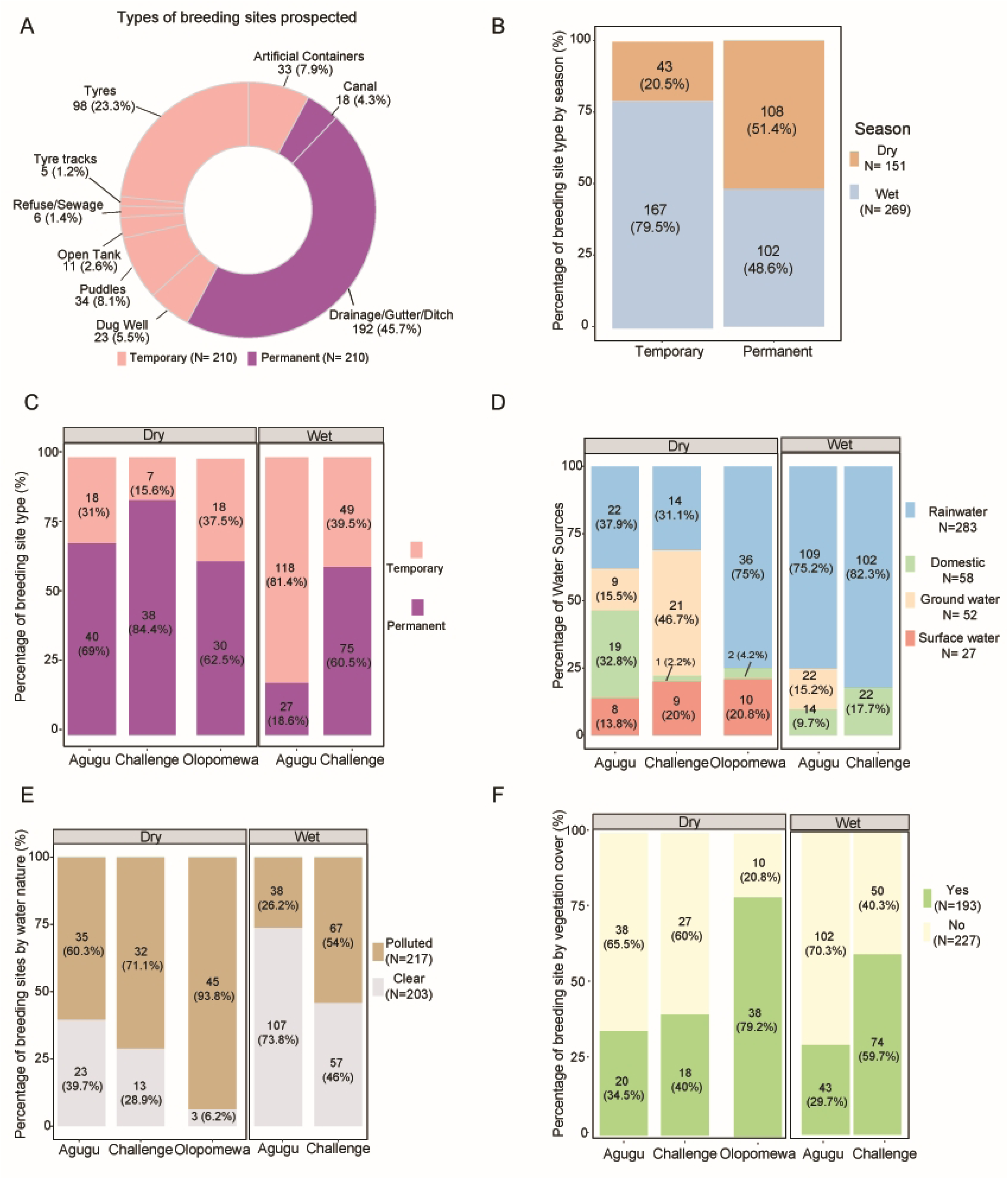
Surveyed potential breeding habitats. (A) Categorized by type (permanent or temporary), (B) By season, (C) By ward and season, (D) By ward and source of water, (E) By ward and water nature, and (F) by ward and vegetation cover.

Overall, potential breeding habitat distribution varied by season and ward. Despite sampling a smaller surface area in Agugu relative to Olopomewa, a slightly higher number of potential breeding habitats were found in Agugu (58/151, 38.4%). Olopomewa has the second highest number of potential breeding sites (48/151, 31.8%) while Challenge has the least (45/151; 29.8%) but also the smallest surface area sampled. During the wet season, larval prospection was conducted only in Agugu and Challenge, where Agugu EAs also contributed a higher proportion of larval habitats (145/269, 53.9%) than Challenge survey locations (124/269, 46.1%) despite the area of the sampled location in Challenge covering nearly four times that of Agugu.

In addition, breeding habitat types also differed across wards and seasons. In the surveyed EAs in Agugu, there was a higher proportion of permanent breeding habitats (40/58, 69.0%) in the dry season, while temporary breeding habitats were more prevalent in the wet season (118/145, 81.4%) (Fig 3C). In contrast, a higher proportion of permanent habitats was observed in Challenge survey locations during both seasons (38/45, 84.4% in the dry season; 75/124, 60.5% in the wet season). Olopomewa was only surveyed during the dry season; therefore, seasonal comparisons were not possible, but majority of the breeding habitats seen were characterized as permanent (30/48, 52.5%) (Fig 3C).

#### Environmental and physicochemical characteristics of potential breeding habitats

Almost all potential breeding habitats were fully exposed to sunlight (412, 98.1%). Rainwater served as the primary water source for those surveyed in Agugu, across both seasons, and in Olopomewa during the dry season (Fig 3D). In Challenge, ground water was the most prevalent water source for surveyed locations during the dry season (21, 46.7%) and rainwater in the wet season (102, 82.3%). Slightly more than half of breeding habitats were polluted (217, 51.6%), and these were more common across all wards during the dry season. In the wet season, the potential breeding habitats identified within surveyed locations in Agugu were mostly clear (107, 73.8%) while polluted ones remained the dominant types in Challenge survey locations (67, 54.0%). (Fig 3E)

Moreover, over half of the breeding habitats (227, 54.4%) had no vegetation cover, a finding that was similar in locations surveyed in Agugu and Challenge during the dry season, whereas most habitats in Olopomewa survey locations (38, 79.2%) had vegetation cover. However, during the wet season, a higher proportion of breeding habitats were covered with vegetation in Challenge survey locations (74, 59.7%) than in Agugu survey locations (43, 29.7%). (Fig 3F). Overall, the pH of potential breeding habitats ranged from 5.0 to 10.0. Mean pH was higher during the dry season than during the wet season (6.86 [SD 0.46] vs 6.71 [1.47]; Kruskal–Wallis χ²=6.01, p=0.014). Across survey locations, mean pH was highest in Challenge (6.86 [1.62]), followed by Agugu (6.69 [0.99]) and Olopomewa (6.61 [0.65]), although no significant differences were observed between locations (Kruskal–Wallis χ²=0.35, p=0.841).

### Assessment of Anopheles larva–positive habitats and molecular identification of collected larvae

#### Anopheles Larva Presence

Among the 420 larval habitats prospected, only 31 (6.7%) had *Anopheles* larvae in them – 5 out of 151 (3.3%) in the dry season and 26 out of 269 (9.7%) in the wet season. Fifteen (48.4%) of the 31 *Anopheles-*positive habitats were permanent and 16 (51.6%) temporary. Most of the larva-positive sites were identified in the wet season (26, 83.9%) as compared to the dry season (5, 16.1%) (Fig 4A). Fig 4A further illustrates that a slightly higher proportion of larva-positive breeding habitats observed in the wet season were temporary as compared to permanent (15, 57.7% vs 11, 42.3%), whereas in the dry season, most breeding habitats with *Anopheles* larvae were permanent (4, 80%).

**Fig 4.**
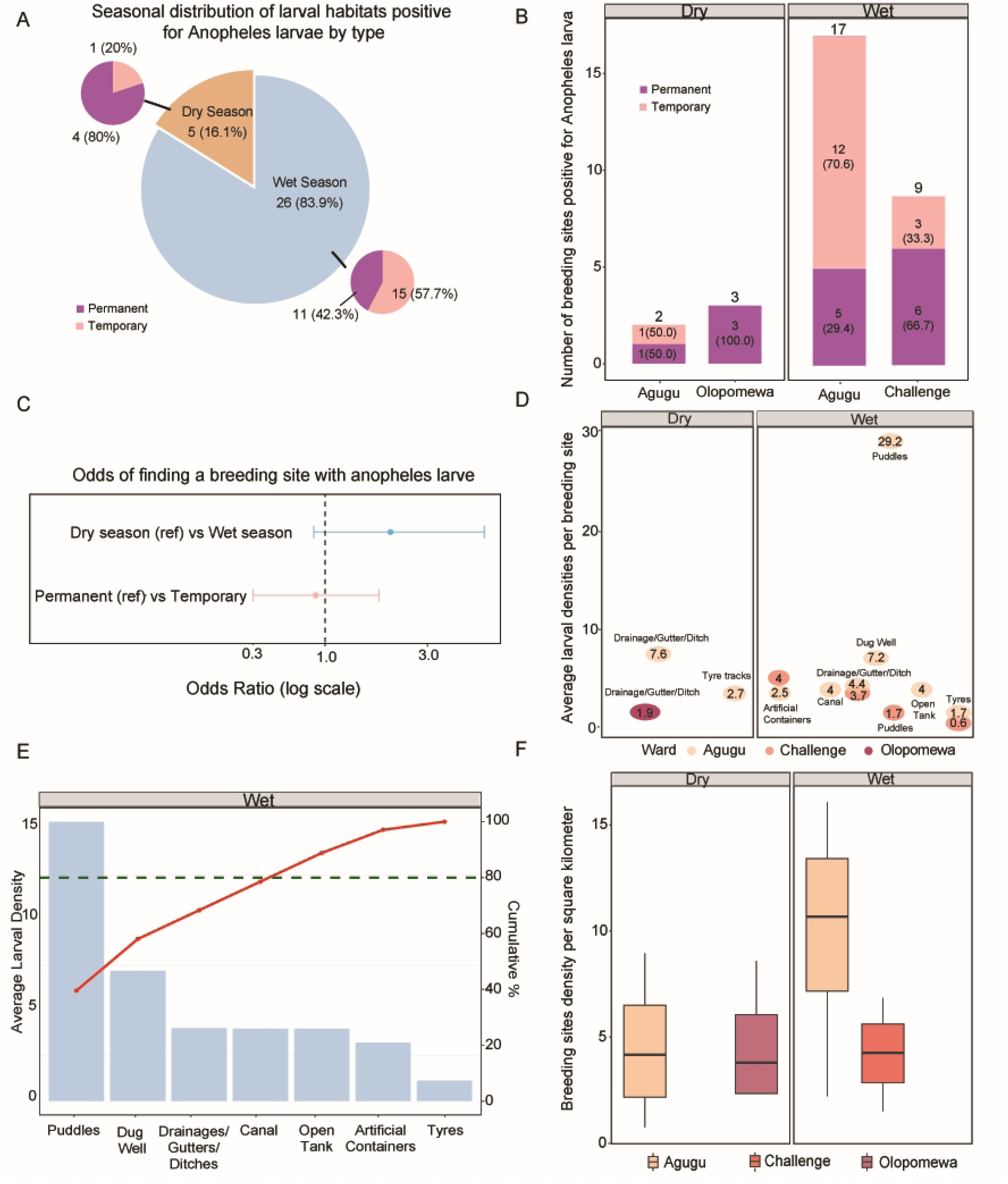
*Anopheles* larval positivity, density, and habitat productivity (A) Seasonal distribution of *Anopheles* larva-positive breeding habitats categorized by permanent and temporary habitat type. (B) Distribution of larva-positive habitats by ward, season and habitat type C) Unadjusted odds ratios for *Anopheles* larval positivity by habitat type (permanent/temporary) and season (D) Average larval densities by breeding habitats type, categorized by ward and season. (E) Pareto analysis of Anopheles larva-positive breeding habitats in the wet season. (F) Anopheles larvae breeding habitats density per km^2^ by ward and season.

The highest numbers of *Anopheles-*positive breeding habitats were found in Agugu’s wet season survey (17/26, 65.4%), whereas most positive habitats in the dry season were found in Olopomewa (3/5, 60.0%). In addition, the composition of these positive habitats differed by breeding site type across wards and seasons. During the dry season, Agugu showed an equal distribution between temporary and permanent habitats (1/2, 50.0% each), whereas all positive habitats identified in Olopomewa were permanent (3/3, 100%). During the wet season, temporary sites accounted for most positive habitats in Agugu (12/17, 70.6%) compared with permanent habitats (5/17, 29.4%), whereas in Challenge, permanent habitats predominated over temporary habitats (6/9, 66.7% vs. 3/9, 33.3%) (Fig 4B).

Overall, larval habitats prospected in the wet season were twice as likely to contain *Anopheles* larva (OR = 2.19, 95% CI: 0.89–6.61). Temporary habitats were found to be less likely to be positive for *Anopheles* larva compared with permanent habitats (OR = 0.89, 95% CI: 0.42–1.87) (Fig 4C). However, none of these associations were statistically significant.

#### Larval density

Average larval density was highest in puddles during the wet season (29.2 larvae per habitat) and in drainages/gutters/ditches during the dry season (7.6 larvae per habitat), and both habitats were found in Agugu. In the wet season, higher mean larval densities were also observed for Agugu compared to Challenge even for the same breeding habitat, except for artificial containers, where densities were higher in Challenge (4.0 vs 2.5 larvae per habitat; Fig 4D).

Pareto analysis further illustrates that across all three ward-level survey locations, puddles, drainages/gutters/ditches, dug wells and canals together accounted for approximately 80% of larval abundance in the wet season (Fig 4E). No statistically significant differences were observed between average larval densities by season, (H = 0.006, p = 0.938) and ward (H = 3.56, p = 0.169) using the Kruskal-Walli’s rank sum test.

#### Molecular Identification

Overall, 1,064 *Anopheles* larvae were collected from the 31 larval-positive habitats, with a higher proportion obtained in the wet season (989, 92.9%) as compared to only 75 (7.1%) during the dry season. Out of the 75 and 989 *Anopheles* larvae collected in the dry and wet season, respectively, only 15 (20%) and 25 (2.5%) of these *Anopheles* larvae were successfully reared to adults. (S1 Fig. panel C). The emerged adult species were mostly *An. gambiae s.s* (12, 86.7%) in the dry season and were all *An. Colluzzi* in the wet season (S1 Fig. panel D)

#### Anopheles-positive larval breeding habitat density estimation

Across 1,000 simulated survey pathways, the estimated density of *Anopheles*-positive breeding habitats per square kilometer differed by ward and season. During the dry season, mean breeding habitat density was relatively low in both wards, with mean of approximately 4 habitats per km² in Olopomewa (mean: 4.41; 95% UI: 1.77-5.83) and 5 habitats per km² in Agugu (mean: 5.09; 95%UI: 0-7.69) surveyed locations. In contrast, during the wet season, breeding habitats density increased substantially in Agugu surveyed locations, with mean densities of approximately 11 habitats per km² (mean: 11.6; 95%UI: 8.59-14.30), while Challenge surveyed locations recorded a mean breeding habitat density of 4.41 habitats per km**²** (95% UI: 2.77–5.83) comparable to dry season values observed in other locations. (Fig 4F)

#### Differences between the environmental and physicochemical characteristics of Anopheles larva-positive and non-positive breeding habitats

The breeding habitats where *Anopheles* larvae were detected were mostly clear in appearance: 69.2% in the wet season (18/26) and 60.0% in the dry season (3/5). Rainfall was the primary water source for most habitats: 84.6% in the wet season (22/26) and 80.0% in the dry season (4/5). An exception was observed in tyre-track habitats during the dry season, where the water originated from wells. However, only 14 habitats (45.1%) were covered with vegetation.

In the dry season, the mean pH in non-positive breeding habitats was slightly higher compared with *Anopheles-*positive larval breeding habitats (6.9 vs 6.7). In contrast, during the wet season, breeding habitats positive for *Anopheles* larvae, had higher mean pH as compared with non-positive breeding habitats (7.0 vs 6.7) (<u>S1 Fig. panel E).</u>

At the ward level, a statistically significant difference in the mean pH was observed in Agugu survey locations during the wet season, with habitats positive for Anopheles larvae having a higher mean pH (7.2) than non-positive breeding habitats (6.5). (p=0.018). However, no significant differences in pH were observed between positive and non-positive habitats within Challenge survey locations in the wet season, nor in either of all the ward-level survey locations during the dry season. <u>(S1 Fig. panel F)</u>

### Spatial relationship between anopheles-positive larval *habitats* and households’ malaria status

Analysis of associations between *Anopheles*-positive larval habitat exposure across dispersal scales and household malaria infections was restricted to households located within the convex hull–defined areas that contained only surveyed habitats for each ward. Following this restriction, the analytic sample comprised 347 individuals from 127 households tested for malaria in Agugu during the dry season and 598 individuals from 218 households during the wet season, with females slightly more represented (53.9%) and approximately 15% of participants under 15 years of age across both seasons. In Challenge, 916 individuals from 344 households were tested during the wet season, while 73 individuals from 35 households located within the entomology study area were tested during the dry season, with a slightly higher proportion of males (64.2%) and approximately 2% under 15 years of age across seasons. In Olopomewa, 156 individuals from 61 households within the surveyed location were tested during the dry season, with a higher proportion of females (58.9%) and approximately 35% under 15 years of age across seasons.

Household malaria positivity varied by ward and season among households located within the defined survey catchment areas. Within these surveyed locations, the highest proportions were observed in Agugu and the lowest in Olopomewa. In Agugu, 21% of sampled households tested positive during the dry season and 18.5% during the wet season. In Challenge, positivity decreased from 8.6% in the dry season to 7.0% in the wet season, while Olopomewa recorded the lowest positivity in the dry season (3.3%) (S5 Appendix).

S2 Fig, panel A presents the AIC profiles across kernel dispersal scales (2m-500m) across wards and seasons. In Challenge, the lowest AIC was recorded at the best fitting scale of approximately 22-24m during the dry season and approximately 30-32m during the wet season. For Agugu survey locations, the best-fitting dispersal was found to be approximately 16-18m during the dry season and 0-2m during the wet season. AIC profiles plateaued beyond approximately 50m across all wards, indicating that model fit did not improve at larger dispersal scales.

Fig 5A shows the kernel-weighted relationship between assumed mosquito dispersal scale and the odds of household malaria positivity across wards and seasons, while the corresponding odds ratios at each ward’s best-fitting dispersal scale are summarised in Fig 5B. Overall, odds ratios were highest at shorter assumed dispersal distances, though the direction and magnitude of associations varied across wards.

**Fig 5.**
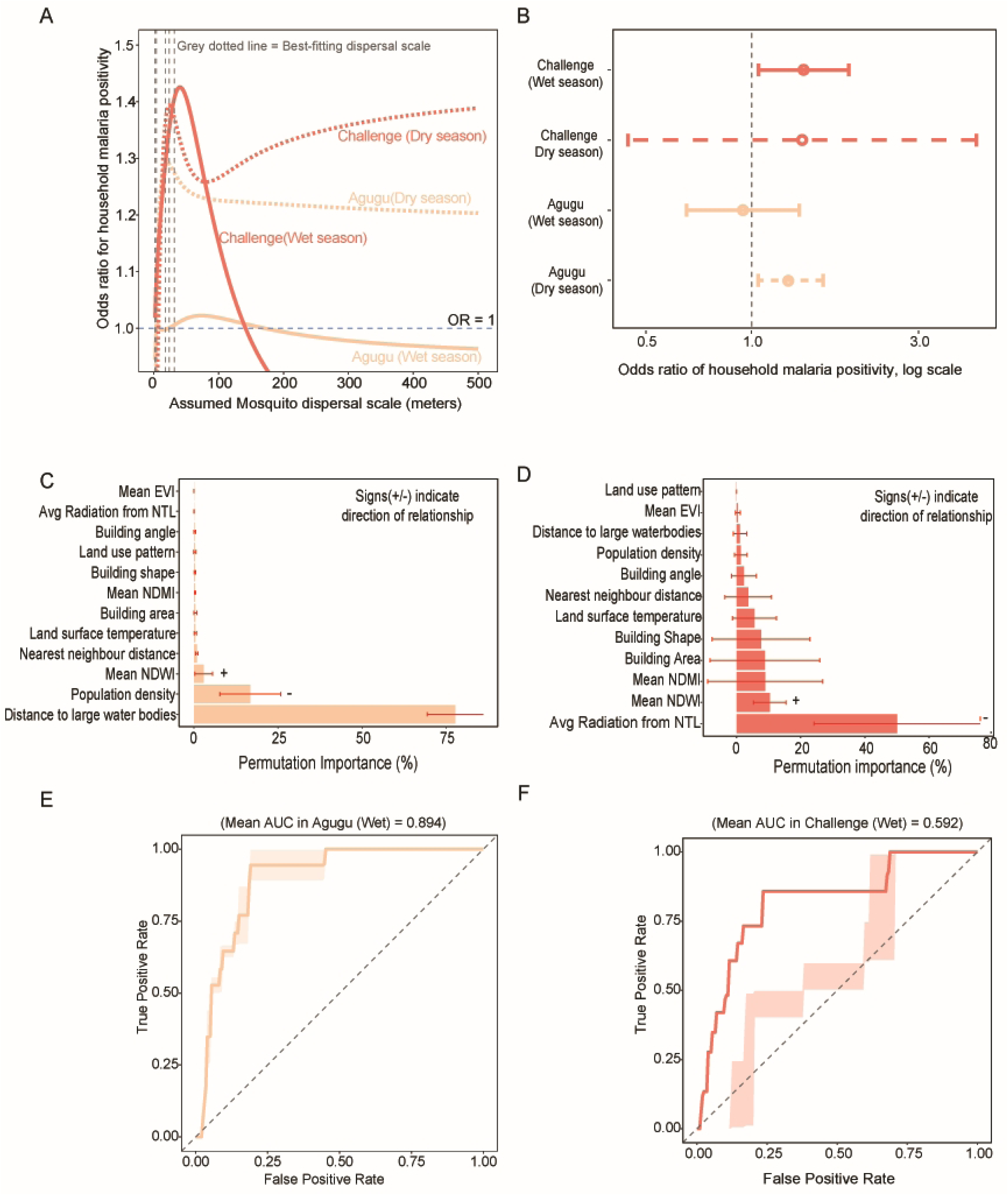
Mosquito dispersal scale, household malaria risk, and environmental drivers of habitat suitability. (A) Relationship between assumed mosquito dispersal scale and household malaria positivity by ward and season. (B) Odd ratios of household malaria positivity at each ward’s best-fitting dispersal scale by season. (C) Bootstrapped permutation importance of environmental predictors in Agugu (wet season). (D) Bootstrapped permutation importance of environmental predictors in Challenge (wet season). (E) Receiver operating characteristic (ROC) curve of MaxEnt models with bootstrapped AUC values in Agugu (wet season). (F) Receiver operating characteristic (ROC) curve of MaxEnt models with bootstrapped AUC values in Challenge (wet season).

In Challenge during the wet season, the OR increased with dispersal distance, peaking near 40m before declining sharply (Fig 5A). At the best-fitting dispersal scale of 30-32m, the OR was statistically significant at 1.41 (95% CI: 1.05–1.89, p = 0.024) (Fig 5B). During the dry season in Challenge, the OR similarly peaked at short dispersal scales before plateauing at larger distances (Fig 5A). At the best-fitting scale of 22-24m, the OR was 1.39 (95% CI: 0.44–4.37, p = 0.569), which was not statistically significant given the wide confidence interval (Fig 5B).

In Agugu during the dry season, the OR showed a modest increase at short dispersal distances, remaining above OR = 1 across larger scales (Fig 5A). At the best-fitting dispersal scale of 16 - 18m, the OR was 1.29 (95% CI: 1.04–1.60, p = 0.019) (Fig 5B). During the wet season in Agugu, the OR remained close to or below OR = 1 across nearly all dispersal scales (Fig 5A), with a non-significant OR of 0.95 (95% CI: 0.65–1.37, p = 0.763) at the best-fitting scale of 0-2m (Fig 5B).

Across all survey locations, the OR for household malaria positivity was highest at short dispersal scales (≤50m); however, the direction and magnitude of the association varied, with only Challenge wet season and Agugu dry season survey locations showing plausible scale-dependent relationships

### Environmental predictors of larval habitat suitability in the wet season

In Agugu surveyed location, predicted habitat suitability in the wet season increased with greater distance from large water bodies, higher population density and positive NDWI values. These variables had the highest permutation importance across bootstrapped model replicates (Fig 5C). Sensitivity analysis showed that distance to large water bodies consistently had the highest permutation importance across models using different temporal averages of vegetation indices, as well as the combined dry-and-wet season model. (S2 Fig, panels B-D)

In Challenge, locations with lower average nighttime light radiation, positive NDWI and negative NDMI values had the highest predicted suitability indices and the highest permutation importance across bootstrapped replicates in the wet season (Fig 5D). Sensitivity analysis also confirmed that lower average nighttime light radiation remained the most influential predictor across models using different temporal averages of vegetation indices. (S2 Fig, panels E-F). S3 Fig presents the response curves of the three strongest environmental predictors in Agugu(A) and Challenge (B) for the wet-season models.

Model performance assessed using the receiver operating characteristic curve, yielded AUC values of 0.894 and 0.592, for Agugu and Challenge wet season models, respectively (Fig 5E–F).

## Discussion

This study presents an assessment of *Anopheles* larval habitats across various urban wards in Ibadan, Nigeria, with implications for vector control policy and practice. The findings suggest that informal settlements embedded within different urban contexts may require different larval source management approaches, as habitat diversity, larval density, the spatial influence of larval habitats on household malaria risk and drivers of habitat suitability varied across surveyed wards. These observations offer insights into the potential value of tailoring larval source management interventions to local ward contexts rather than adopting a uniform city-wide approach and may assist the National Malaria Elimination Programme (NMEP) in refining larval source management strategies particularly larviciding while enhancing malaria control efforts in urban settings. Further studies, however, are needed to confirm these findings prior to policy adoption.

### Breeding habitats are findable

Our findings demonstrate the presence of diverse larval habitats in urban areas, with the equal distribution of permanent and temporary breeding habitats reflecting the heterogeneous nature of urban landscapes. The potential *Anopheles* larval breeding habitats identified, included drainages, gutters, ditches, puddles, tyres, tyre tracks, and artificial containers such as plastic bowls and earthen pots, the majority of which were anthropogenic in origin. This finding aligns with studies in urban Nigeria, where both permanent and temporary breeding habitats contribute to a heterogeneous and complex larval ecology. [10, 35–37] Similar patterns have also been documented within the last decade across other urban African settings such as Yaoundé, Accra, Cotonou, Benin and Dar es Salaam, which documented human-made larval habitats such as puddles, drains, and construction sites as breeding habitats. [38–42] These findings further confirm the capacity of *Anopheles* species to thrive in diverse urban environments, (plausibly due to urbanization) and emphasize the need for comprehensive, urban-specific larval source management strategies.

### Seasonal Variability

Furthermore, we found that during the dry season, permanent breeding habitats including drainages, gutters, and ditches dominate as breeding habitats, particularly within the surveyed locations in Olopomewa. In contrast, during the wet season, a wider variety of habitats such as artificial containers, tyres and puddles, become breeding habitats in addition to the ones found in the dry season. This pattern of seasonal variation in *Anopheles* larval habitats is in consonance with studies in other parts of Southwest Nigeria. In Akure North, Ondo State, Olusi et al. (2021) observed that during the dry season, *Anopheles* larvae predominantly inhabited dirty or muddy environments such as gutters, ditches, and canals, while the rainy season expanded the range of breeding habitats to include abandoned tyres, potholes, and rain pools.[43] Similarly, a study in Osun State identified ground pools, gutters, containers, and abandoned tyres as key larval habitats, with ground pools being more prominent during the rainy season. [9] However, some studies in Northern Nigeria, reported that *Anopheles* breeding during the dry season was documented in unconventional sites such as polluted open sewers, swamps and areas contaminated with human waste [44–45]. This may have resulted from the prolonged dry seasons in Northern Nigeria, which reduce the availability of typical breeding habitats and drive *Anopheles* mosquitoes to exploit unconventional habitats, reflecting their adaptability to changing ecological and climatic conditions. Furthermore, Ndiaye et al, in their study in 3 urban districts in Senegal reported the seasonal variation in the number of *Anopheles* breeding habitats which decreased during the dry season when more temporary breeding habitats dried out. [46] These findings demonstrate the seasonal selection in breeding habitats by *Anopheles* larva, and the increased larval abundant breeding habitats during the wet season has important implications for the timing of LSM particularly larviciding.

### Settlement-Variability in Anopheles Breeding Habitats

Our study findings also showed that a diverse and majority of positive breeding habitats were anthropogenic and seen in surveyed locations in Agugu-a ward predominated by slum settlements, whereas Olopomewa and Challenge survey locations had fewer types of breeding habitats with *Anopheles*’ larva. This finding is in consonance with previous studies conducted in urban slums, which identified anthropogenic temporary breeding habitats such as ground pools and discarded containers as the predominant larval habitats. [35,42] This is, however, not unexpected as slum settlements by definition are areas with poor housing and waste management infrastructure hence anopheles breeding habitats are more likely to thrive in such environments. Strengthening environmental sanitation in these areas could curtail these temporary breeding habitats thereby limiting potential *Anopheles* breeding habitats and lessening the need for extensive larviciding interventions in urban slums. In addition, the presence of fewer and more predictable breeding habitat types — such as drainages, gutters, ditches, and artificial containers — dominating in the formal and informal settlement-predominated wards - across both seasons poses an opportunity to streamline larval source management efforts to these breeding habitats in such wards and potentially lower operational costs.

### Seasonal and Settlement Patterns of Anopheles Larval Density

Quantifying *Anopheles* larval densities is essential for identifying high-yield breeding habitats, enabling larval source management efforts to be focused on where they may have the greatest impact. In our study, Agugu surveyed catchment area had the highest larval densities in both seasons as compared to Olopomewa in the dry season and Challenge in the wet season. Further analysis revealed that puddles had the highest larval density in the wet season while gutters/drainages and canals had the highest in the dry season. Busari et al., in their study in an urban community in Osun State reported significantly higher larva density in ground pools during the wet season, a finding consistent with our study. [9] Likewise, Oforka et al, observed that permanent breeding habitats such as gutters had significantly greater larval densities than temporary habitats within slum communities particularly during the dry season.[12] These variations in larval density pattern across habitat types and seasons suggest the need for seasonal and settlement adaptive larval source management (LSM) strategies.

### Spatial Relationship Between Larval Habitats and Malaria Risk

Our analysis of the spatial relationship between *Anopheles*-positive larval habitats and household malaria status revealed that households closer to these habitats experienced higher malaria risk as documented by previous studies [47–48]. However, within the surveyed catchment area of Challenge, an informal-settlement-dominant ward, this risk was largely confined to shorter distances from households, suggesting the presence of localized breeding clusters. In contrast, within the surveyed catchment area of Agugu, malaria risk persisted at greater distances, indicating the presence of potentially more dispersed and widespread breeding habitats or additional drivers of transmission risk. These findings support the growing recognition that malaria risk within urban environments is often highly localized rather than uniformly distributed. The differing risk gradients observed across study wards suggest that settlement archetype may influence not only larval habitat distribution but also the spatial scale at which malaria control interventions are likely to be most effective.

### Habitat distribution and suitability

In our habitat suitability analysis, we found that almost all parts of the wards were suitable for *Anopheles* larvae with the highest suitability associated with increasing distance from large water bodies, areas with higher population density in Agugu and areas with lower nighttime lights in Challenge. The finding that suitability increased with distance from large water bodies suggests that breeding is more concentrated in smaller, stagnant, human-made water collections—such as puddles, artificial containers, receptable—rather than in natural large water bodies. This could explain our findings where we found puddles having the highest larval densities in Agugu surveyed locations. Moreover, in Challenge survey locations, areas with lower nighttime illumination —likely representing less densely built or poorly lit peripheral areas—showed higher suitability, suggesting that even within informal-settlement-predominated wards, peripheral, overlooked water collection areas or poorly serviced zones may support residual breeding of *Anopheles*. These findings are consistent with literature in some other parts of Africa which reported proximity of larval habitats to household’s water bodies and light intensity as drivers of *Anopheles* larval breeding [49–51]. In addition, we found that vegetation indices, especially NDWI, were main predictors of suitability. Though, other studies in Nigeria have reported mainly climatic factors like rainfall, precipitation and temperature as predictors of habitat suitability in contrast to our study findings, these studies were conducted at larger spatial scales i.e. national, state or local government where climatic variability is greater. [52–53] Our study, however, used a more granular area (ward level) which could explain the differences in the covariates available for prediction.

### Emerging Considerations for Urban Larval Source Management

The presence of *Anopheles* larvae in a variety of breeding habitats in urban areas—ranging from drainages to puddles, tyres, and tyre tracks—highlights the adaptability of these vectors within urban environments. The finding that a substantial proportion of breeding habitats harbored *Anopheles* larvae in the urban areas, ranging approximately 10% drainages/gutters/ditches during the wet season to 1 out of 2 tyre tracks in the dry season, suggests that urban areas may sustain conditions conducive to malaria vector breeding. However, given the exploratory nature of the study and the sampling approach employed, these estimates should be interpreted with caution. Nevertheless, the relatively low breeding habitat density per square kilometer in both the urban slum and formal or informal-settlement-dominant wards indicates that these habitats are relatively few, potentially creating opportunities to allocate resources efficiently and targeted operational planning while focusing on high-priority habitats, although further studies are needed to determine whether these observations reflect broader urban patterns or characteristics of the wards studied.

While urban larval habitats in surveyed catchment area of Agugu are heterogeneous, their predictability in certain sites—such as puddles, tyres, and tyre tracks—supports the inclusion of environmental management as a key component of larval source management strategies in urban slums. Similarly, the persistence of malaria risk over greater distances from larval habitats suggests breeding habitats may be more disperse or cryptic, raising the possibility that larviciding alone may be insufficient where other environmental exposures contribute to risk. Understanding whether this pattern reflects differences in habitat distribution, unmeasured environmental exposures, or other drivers of malaria risk will require further investigation.

In the informal- and formal-settlement-predominated ward, drainages, gutters, and canals accounted for a large proportion of productive habitats across both wet and dry seasons. While this pattern may point to the potential importance of routine drainage maintenance within integrated larval source management strategies, the study design does not allow definitive conclusions regarding the effectiveness of specific interventions. Additionally, the observed differences could motivate further investigation into whether prioritizing such habitats during larviciding might improve the efficiency of LSM in such settlement archetypes

Furthermore, the more localized relationship observed between household malaria risk and larval habitats in the informal-settlement-dominated ward provides an opportunity to evaluate whether focal targeting of larval source management interventions around households could be effective. Conversely, the implications of the broader spatial extent of malaria risk observed in the slum-dominated ward for spatial coverage of larval source management interventions remain unclear and may require further investigation.

Additionally, this study demonstrates the feasibility of using remote-sensed earth observation and other socio-environmental data to explore the suitability of habitats for anopheles breeding at the lowest administrative level in Nigeria. Further research is needed to determine whether integrating these data into larval source management planning improves the targeting, operational efficiency, and cost-effectiveness of larviciding and other larval source management activities in urban areas, and whether such gains can be achieved under routine programme conditions.

While this study provides insights into larval source management in urban landscapes, some limitations should be acknowledged. First, survey locations were selected through a community-led process informed by preliminary data rather than a fully systematic scientific sampling design, largely due to resource constraints which could have resulted in the small number of habitats prospected. This may limit the generalizability of the findings, particularly regarding the spatial distribution and density of larva-positive breeding habitats across surveyed wards. Although a simulated pathway sampling approach was used to enhance coverage, some inaccessible breeding habitats may have been missed, potentially resulting in an underestimation of true breeding habitat density. Second, the distance–decay analysis evaluating the relationship between household malaria risk and larvae-positive habitats, relied on aggregate household malaria risk without accounting for individual-level behavioural factors. In addition, epidemiological and entomological data were not collected concurrently, although both were collected during the same season, which constrains causal interpretation of the observed associations. Nevertheless, we assumed that productive breeding habitats, particularly permanent habitats, tend to persist across seasons and recur in similar locations over time, so the observed spatial patterns can still be considered informative, albeit hypothesis-generating. Thirdly, restricted access to environmental and climatic data at the scale of the study area may have affected the performance of our habitat suitability models. In addition, only a single ward representing each settlement archetype was included.

Consequently, observed differences may reflect characteristics of the specific wards studied rather than settlement archetype alone, and findings should be interpreted as hypothesis-generating. Despite these limitations, this study is among the few to provide preliminary programmatic insights into larval ecology, contribution to household malaria risk and spatial heterogeneity across different urban settlement types at the lowest administrative level (wards) in Nigeria, the findings may help inform future ward-level surveillance, microplanning, and evaluation of larval source management strategies in urban Nigeria.

## Conclusion

This exploratory study provides preliminary evidence that differences in larval habitat characteristics, habitat suitability, and the spatial influence of larval habitats on malaria risk may exist across informal settlements embedded within different urban wards. These differences have potential implications for how larval source management interventions are planned and targeted. The study further demonstrates the feasibility of integrating entomological, epidemiological, and geospatial data to characterize urban larval ecology at the ward level. Further research across a larger number of wards and urban settings is needed to determine whether the patterns observed in this study are reproducible and sufficiently consistent to inform larval source management planning under routine programme conditions.

## Data Availability

The minimal data set and accompanying analysis code will be made publicly available without restriction in Harvard Dataverse upon acceptance. The DOI and public access link will be provided before publication.

## Acknowledgement

We thank the residents and community leaders of the selected wards for their cooperation and support during field activities. We are grateful to the local community guides and field teams who assisted with household surveys, larval habitat identification, and larval sampling, as well as the laboratory and data management team. We also acknowledge the support of relevant stakeholders who facilitated community entry, multi-stakeholders dialogue and implementation of the field study.

## Supporting information

S1 Table. Operational definitions of larval breeding habitat classifications used in the field survey.

S1 Appendix. Distribution of settlement composition by ward.

S2 Appendix. Example output from the simulated pathway analysis used to estimate breeding habitat density in Agugu during the wet season.

S2 Table. Environmental predictor variables, source datasets, and acquisition years. S3 Appendix. Correlation matrix of environmental predictors in Agugu.

S4 Appendix. Correlation matrix of environmental predictors in Challenge.

S1 Fig. Seasonal, molecular, and physicochemical characteristics of Anopheles larval breeding habitats. (A) Seasonal breakdown of breeding habitat types during the dry season. (B) Seasonal breakdown of breeding habitat types during the wet season. (C) Number of Anopheles larvae successfully reared to adults, by season. (D) Anopheles species composition, by season. (E) Mean pH of Anopheles-positive versus non-positive breeding habitats, by season. (F) Mean pH of Anopheles-positive versus non-positive breeding habitats by ward and season.

S5 Appendix. Household malaria positivity by ward and season.

S2 Fig. Sensitivity analysis of AIC profiles and environmental predictor importance across temporal averages. (A) AIC profiles across kernel dispersal scales (0m–40m), by ward and season. (B–D) Sensitivity analysis of permutation importance for environmental predictors in Agugu using different temporal averages of vegetation indices and the combined dry-and-wet season model. (E–F) Sensitivity analysis of permutation importance for environmental predictors in Challenge using different temporal averages of vegetation indices.

S3 Fig. Response curves of the three strongest environmental predictors of Anopheles larval habitat suitability in the wet season. (A) Agugu. (B) Challenge.

